# Detection of dementia on raw voice recordings using deep learning: A Framingham Heart Study

**DOI:** 10.1101/2021.03.04.21252582

**Authors:** Chonghua Xue, Cody Karjadi, Ioannis Ch. Paschalidis, Rhoda Au, Vijaya B. Kolachalama

## Abstract

**Background:** Identification of reliable, affordable, and easy-to-use strategies for detection of dementia are sorely needed. Digital technologies, such as individual voice recordings, offer an attractive modality to assess cognition but methods that could automatically analyze such data are not readily available.

**Methods and findings:** We used 1264 digital voice recordings of neuropsychological examinations administered to participants from the Framingham Heart Study (FHS), a community-based longitudinal observational study. The recordings were 73 minutes in duration, on average, and contained at least two speakers (participant and clinician). Of the total voice recordings, 483 were of participants with normal cognition (NC), 451 recordings were of participants with mild cognitive impairment (MCI), and 330 were of participants with dementia (DE). We developed two deep learning models (a two-level long short-term memory (LSTM) network and a convolutional neural network (CNN)), which used the raw audio recordings to classify if the recording included a participant with only NC or only DE and to differentiate between recordings corresponding to those that were non-demented (NDE (NC+MCI)) and DE. Based on 5-fold cross-validation, the LSTM model achieved a mean (±std) area under the receiver operating characteristic curve (AUC) of 0.740±0.017, mean balanced accuracy of 0.647±0.027, and mean weighted F1-score of 0.596±0.047 in predicting cases with DE from those with NC. The CNN model achieved a mean AUC of 0.805±0.027, mean balanced accuracy of 0.743±0.015, and mean weighted F1-score of 0.742±0.033 in predicting cases with DE from those with NC. For the task related to classification of participants with DE from NDE, the LSTM model achieved a mean AUC of 0.734±0.014, mean balanced accuracy of 0.675±0.013, and mean weighted F1-score of 0.671±0.015. The CNN model achieved a mean AUC of 0.746±0.021, mean balanced accuracy of 0.652±0.020, and mean weighted F1-score of 0.635±0.031 in predicting cases with DE from those who were NDE.

**Conclusion:** This proof-of-concept study demonstrates the potential that raw audio recordings of neuropsychological testing performed on individuals recruited within a community cohort setting can facilitate dementia screening.

## Introduction

Impairment in cognition is a common manifestation among all the individuals suffering from dementia, of which Alzheimer’s disease (AD) is the most common. Despite the rising dementia epidemic accompanying a rapidly aging population, there remains a paucity of cognitive assessment tools that are applicable regardless of age, education, and language/culture. Thus, there is an urgent need to identify reliable, affordable, and easy-to-use strategies for detection of signs of dementia. Starting in 2005, the Framingham Heart Study (FHS) began digitally recording all spoken responses to neuropsychological tests. These digital voice recordings allow for precise capture of all responses for verbal tests and a novel application of the Boston Process Approach (BPA) [1]; a scoring method that emphasizes how a participant makes errors to differentiate between participants with similar test score results. With the emergence of speech recognition and analysis tools, there was a realization that these recordings used for quality control were now data in of themselves because speaking is a highly complex cognitive skill. Virtually all voice recognition software requires speech-to-text transcription from which to extract linguistic measures of speech and manual transcription is often required to reach high levels of accuracy. This manual work is not only tedious, but also to ensure high levels of accuracy often requires some level of training in speech-to-text transcription and quality control to document transcription accuracy. Such expertise is not readily available at all locations around the globe. Developing a computational tool that can simply take a raw voice recording as an input and automatically assess the dementia status of the individual has broad implications for dementia screening tools that are scalable across diverse populations.

Promising solutions to tackle such datasets may be found in the field of deep learning, an approach to data analysis that has increasingly been used over the past few years to address an array of formerly intractable questions in medicine. Deep learning [2], a subfield of machine learning, is based upon specific models known as neural networks which decompose the complexities of observed datasets into hierarchical interactions among low-level input features. Once these interactions are learned, refined, and formalized by exposure to a training dataset, fully trained deep learning models leverage their “experience” of prior data to make predictions about new cases. Thus, these approaches offer powerful medical decision-making potential due to their ability to rapidly identify low-level signatures of disease from large datasets, and quickly apply them at scale. This hierarchical approach makes deep learning ideally suited to derive novel insights from high volumes of audio/voice data. Our primary objective was to develop a long short-term memory network (LSTM) model and a convolutional neural network (CNN) model, to predict dementia status on the FHS raw voice recordings, without manual feature processing of the audio content. As a secondary objective, we processed the model-derived salient features and reported the distribution of time spent by individuals on various neuropsychological tests and these tests’ relative contributions to dementia assessment.

## Methods

### Study population

The voice data consists of digitally recorded neuropsychological examinations administered to participants from the Framingham Heart Study (FHS), which is a community-based longitudinal observational study that was initiated in 1948 and consists of several generations of participants [3]. FHS began in 1948 with the initial recruitment of the Original Cohort (Gen 1), added the Offspring Cohort in 1971 (Gen 2), the Omni Cohort in 1994 (OmniGen 1), a Third Generation Cohort in 2002 (Gen 3), a New Offspring Spouse Cohort in 2003 (NOS), and a Second Generation Omni Cohort in 2003 (OmniGen 2). The neuropsychological examinations consist of multiple tests that assess memory, attention, executive function, language, reasoning, visuoperceptual skills, and premorbid intelligence. Further information and details can be found in Au et. al [4], including the lists of all the neuropsychological tests included in each iteration of the FHS neuropsychological battery [5].

### Cognitive status determination

The cognitive status of the participants over time was diagnosed via the FHS dementia diagnostic review panel [6, 7]. The panel consists of at least one neuropsychologist and at least one neurologist. The panel reviews neuropsychological and neurological exams, medical records, and family interviews for each participant. Selection for dementia review is based on whether participants have shown evidence of cognitive decline, as has been previously described [4].

The panel creates a cognitive timeline for each participant that provides the participant’s cognitive status on a given date over time. To label the participants’ cognitive statuses at the time of each recording, we selected the closest date of diagnosis to the recording that fell on or before the date of recording or after the recording within 180 days. If the closest date of diagnosis was more than 180 days after the recording, but the participant was determined to be cognitively normal on that date, we labeled that participant as cognitively normal. Dementia diagnosis was based on criteria from the *Diagnostic and Statistical Manual of Mental Disorders*, fourth edition (DSM-IV) and the NINCDS-ADRDA for Alzheimer’s dementia [8]. The study was approved by the Institutional Review Boards of Boston University Medical Center and all participants provided written consent.

### Digital voice recordings

FHS began to digitally record the audio of neuropsychological examinations in 2005. Overall, FHS has 9786 digital voice recordings from 5449 participants. It must be noted that not all participants underwent dementia review and repeat recordings were available on some participants. For this study, we selected only those participants who underwent dementia review, and their cognitive status was available (normal/MCI/dementia). The details of how participants are flagged for dementia review has been previously described [9]. In total, we obtained 656 participants with 1264 recordings, which are 73.13 (±22.96) minutes in duration on average. The range is from 8.43 to 210.82 minutes. There are 483 recordings of participants with normal cognition (NC; 291 participants), 451 recordings of participants with mild cognitive impairment (MCI) (309 participants), 934 recordings of non-demented participants (NDE; 507 participants), and 330 recordings of participants with dementia (DE; 223 participants) **(Tables 1 & S1)**. Of the 656 participants, one participant may have several recordings with different cognitive statuses. For example, one participant could have NC at their first recording, MCI at their second recording, and DE at their third recording. This implies that participants with a recording associated with either NC, MCI, or DE are not mutually exclusive and will not necessarily add up to the overall 656 participants. The recordings were obtained in the WAV format and downsampled to 8 kHz.

**Table 1:** Demographics and participant characteristics. For each participant, digital voice recordings of neuropsychological examinations were collected. Tables (A), (B) and (C) show the demographics of the participants with normal cognition, mild cognitive impairment, and dementia, respectively at the time of the voice recordings. Here, N represents the number of participants. The mean age (± standard deviation) is reported at the time of the recordings. Mean MMSE scores (± standard deviation) were computed closest to the time of the voice recording. For cognitively normal participants, ApoE data was unavailable for one Generation 1 (Gen 1) participant, and eight Generation (Gen) 2 participants; MMSE data was not collected on Generation (Gen) 3 participants. For MCI participants, ApoE data was unavailable for one Gen 1 participant, six Gen 2 participants, and one New Offspring Cohort (NOS) participant; MMSE data was also not collected for OmniGen2, and NOS participants and was not available for one Gen 1 participant. For demented participants, ApoE data was unavailable for six Gen 1 participants and three Gen 2 participants; MMSE data was not collected for Gen 3, OmniGen2, and NOS Cohort participants and not available for one Gen 1 participant.

| **Normal cases** |  |  |  |  |  |  |
| --- | --- | --- | --- | --- | --- | --- |
| **Cohort** | **N** | **Female** | **ApoE4+** | **Recordings** | **Age (years)** | **Mean MMSE** |
| Gen 1 | 42 | 28 | 6 | 75 | 90.9±3.0 | 27.5±2.0 |
| Gen 2 | 238 | 117 | 40 | 392 | 73.9±7.8 | 28.1±2.0 |
| Gen 3 | 4 | 1 | 0 | 7 | 60.4±11.5 | NA |
| OmniGen 1 | 7 | 2 | 2 | 9 | 71.4±9.3 | 26.6±2.6 |
| Total | 291 | 148 | 48 | 483 | 76.3±9.8 | 27.9±2.0 |

**(B)**
| **MCI cases** |  |  |  |  |  |  |
| --- | --- | --- | --- | --- | --- | --- |
| **Cohort** | **N** | **Female** | **ApoE4+** | **Recordings** | **Age (years)** | **Mean MMSE** |
| Gen 1 | 64 | 47 | 13 | 85 | 91.4±3.1 | 26.0±2.4 |
| Gen 2 | 235 | 124 | 67 | 353 | 79.3±6.8 | 27.2±2.2 |
| Gen 3 | 1 | 0 | 0 | 1 | 60.0±0.0 | NA |
| OmniGen 1 | 6 | 1 | 1 | 7 | 73.1±6.8 | 25.3±2.3 |
| OmniGen 2 | 1 | 1 | 1 | 1 | 74.0±0.0 | NA |
| NOS | 2 | 1 | 0 | 4 | 86.5±5.3 | NA |
| Total | 309 | 267 | 82 | 451 | 81.5±8.0 | 26.9±2.3 |

(C)
| **Dementia cases** |  |  |  |  |  |  |
| --- | --- | --- | --- | --- | --- | --- |
| **Cohort** | **N** | **Female** | **ApoE4+** | **Recordings** | **Age**<br>**(years)** | **Mean**<br>**MMSE** |
| Gen 1 | 78 | 56 | 16 | 99 | 92.2±3.1 | 21.3±5.7 |
| Gen 2 | 139 | 84 | 40 | 224 | 82.1±6.7 | 23.3±5.5 |
| Gen 3 | 1 | 0 | 0 | 1 | 80.0±0.0 | NA |
| OmniGen 1 | 3 | 1 | 0 | 4 | 71.5±1.7 | 24.8±1.5 |
| OmniGen 2 | 1 | 1 | 1 | 1 | 77.0±0.0 | NA |
| NOS | 1 | 1 | 0 | 1 | 80.0±0.0 | NA |
| Total | 223 | 143 | 57 | 330 | 84.9±7.6 | 22.7±5.6 |

We observed that the FHS participants on average spent different amounts of time to complete specific neuropsychological tests, and these times varied as a function of their cognitive status for most of the tests (**Figure 1**). For example, almost all participants spent the highest amount of time while completing the Boston Naming Test (BNT). During this test, participants who were DE spent significantly more time (611.1±260.2 s) than the participants who are not demented (NDE; 390.7±167.4 s). This observation was also valid when the time spent by participants with DE was compared specifically with those with NC (405.9±176.8 s), or with the ones who had MCI (321.2±93.8 s). However, there was no statistical significance between the times taken for BNT by participants with NC and MCI. A similar pattern was observed for a few other neuropsychological tests including ‘Visual Reproductions Delayed Recall’, ‘Verbal Paired Associates Recognition’, ‘Copy Clock’, ‘Trails A’, and ‘Trails B’ (**Table 2**). We also observed no statistically significant differences in the times taken for the participants with NC, MCI, DE and NDE on a few other neuropsychological tests including ‘Logical Memory Immediate Recall’, ‘Digit Span Forward’, ‘Similarities’, ‘Verbal Fluency (FAS)’, ‘Finger Tapping’, ‘Information (WAIS-R)’, and ‘Cookie Theft’ (**Table 2**).

**Table 2:** Time spent on the neuropsychological tests. Average time spent (± standard deviation) by the FHS participants on each neuropsychological test is shown. For each test, average values (± standard deviation) were computed on participants with normal cognition (NC), those with mild cognitive impairment (MCI), and those who had dementia (DE); the no dementia group (NDE) combined the NC and MCI individuals. All reported time values are in minutes. Logical memory (LM) tests with a (†) symbol denote that an alternative story prompt as administered for the test. It is possible that one participant may receive a prompt under each of the LM recall conditions (one recording).

|  | NC | MCI | NDE | DE |
| --- | --- | --- | --- | --- |
| **Demographics** | 244.4±151.0 | 178.7±106.4 | 231.0±144.4 | 295.5±163.2 |
| **Logical Memory Immediate Recall** | 135.0±28.6 | 116.9±19.5 | 131.6±27.8 | 128.9±38.5 |
| **Logical Memory Immediate Recall (†)** | | | | 123.0±19.3 |
| **Visual Reproductions Immediate Recall** | 219.3±79.2 | 190.8±37.9 | 213.9±73.8 | 236.1±67.9 |
| **Verbal Paired Associates Learning** | 367.6±80.4 | 366.4±65.3 | 367.4±77.3 | 414.3±155.1 |
| **Digit Span Forward** | 115.9±31.4 | 100.1±32.3 | 113.0±31.9 | 107.5±36.7 |
| **Digit Span Backward** | 109.0±42.5 | 128.2±44.7 | 112.6±43.1 | 132.6±52.6 |
| **Logical Memory Delayed Recall** | 86.3±37.8 | 57.1±20.2 | 80.8±36.8 | 54.0±25.3 |
| **Logical Memory Delayed Recall (†)** | | | | 33.0±7.1 |
| **Logical Memory Multiple Choice** | 89.4±18.3 | 103.8±31.2 | 91.9±21.4 | 138.9±62.3 |
| **Logical Memory Multiple Choice (†)** | | | | 159.5±14.8 |
| **Visual Reproductions Delayed Recall** | 140.6±73.2 | 114.8±28.3 | 136.0±67.9 | 79.1±48.2 |
| **Visual Reproductions Multiple Choice** | 63.5±26.5 | 60.9±27.8 | 63.0±26.5 | 77.5±37.1 |
| **Verbal Paired Associates Recall** | 66.2±23.5 | 85.6±43.7 | 69.5±28.4 | 90.3±48.7 |
| **Verbal Paired Associates Recognition** | 75.8±21.6 | 90.0±26.4 | 78.3±22.9 | 126.0±52.2 |
| **Similarities** | 227.8±89.6 | 217.3±79.5 | 225.9±87.3 | 228.0±120.4 |
| **Command Clock** | 80.9±31.2 | 83.3±29.1 | 81.3±30.5 | 133.0±82.2 |
| **Verbal Fluency** | 325.5±39.0 | 330.0±29.2 | 326.3±37.2 | 336.1±65.4 |
| **Boston Naming Test** | 405.9±176.8 | 321.2±93.8 | 390.7±167.4 | 611.1±260.2 |
| **Copy Clock** | 70.1±33.5 | 54.7±14.0 | 67.0±31.1 | 90.9±44.4 |
| **Trails A** | 115.4±58.3 | 105.3±23.7 | 113.5±53.3 | 199.0±107.1 |
| **Trails B** | 219.2±107.6 | 241.0±117.5 | 223.4±108.5 | 431.2±278.0 |
| **WRAT-3 Reading** | 119.7±36.8 | 115.1±52.3 | 118.8±39.7 | 142.8±70.4 |
| **Hooper Visual Organization Test** | 398.5±182.1 | 284.1±90.8 | 376.6±173.7 | 424.6±187.4 |
| **Block Design (WAIS)** | 357.3±213.6 | 389.7±145.0 | 360.9±205.1 | 487.3±214.3 |
| **Finger Tapping** | 294.4±106.9 | 323.8±87.6 | 300.3±102.3 | 289.0±130.6 |
| **Information (WAIS-R)** | 392.7±181.2 | 335.8±156.9 | 385.1±176.7 | 372.7±153.6 |
| **Cookie Theft** | 198.9±75.3 | 244.6±101.7 | 206.5±79.6 | 186.9±147.0 |
| **Digit Symbol Coding** | 204.6±76.1 |  | 204.6±76.1 | 269.5±116.2 |
| **Digit Symbol Learning** | 125.0±75.7 |  | 125.0±75.7 | 90.6±78.7 |
| **Digit Symbol Recall** | 67.4±29.8 |  | 67.4±29.8 | 91.4±122.7 |
| **Clock Drawing Number Placement** | 47.6±40.6 | 51.8±20.5 | 48.5±37.2 | 81.6±71.5 |
| **Clock Drawing Time Setting** | 44.0±26.9 | 37.9±7.6 | 42.8±24.5 | 75.3±59.3 |
| **Math Fluency** | 53.1±66.0 |  | 53.1±66.0 | 187.4±84.2 |
| **Balance Physical Function Test** | 341.2±80.3 |  | 341.2±80.3 | 202.5±71.4 |

**Figure 1:**
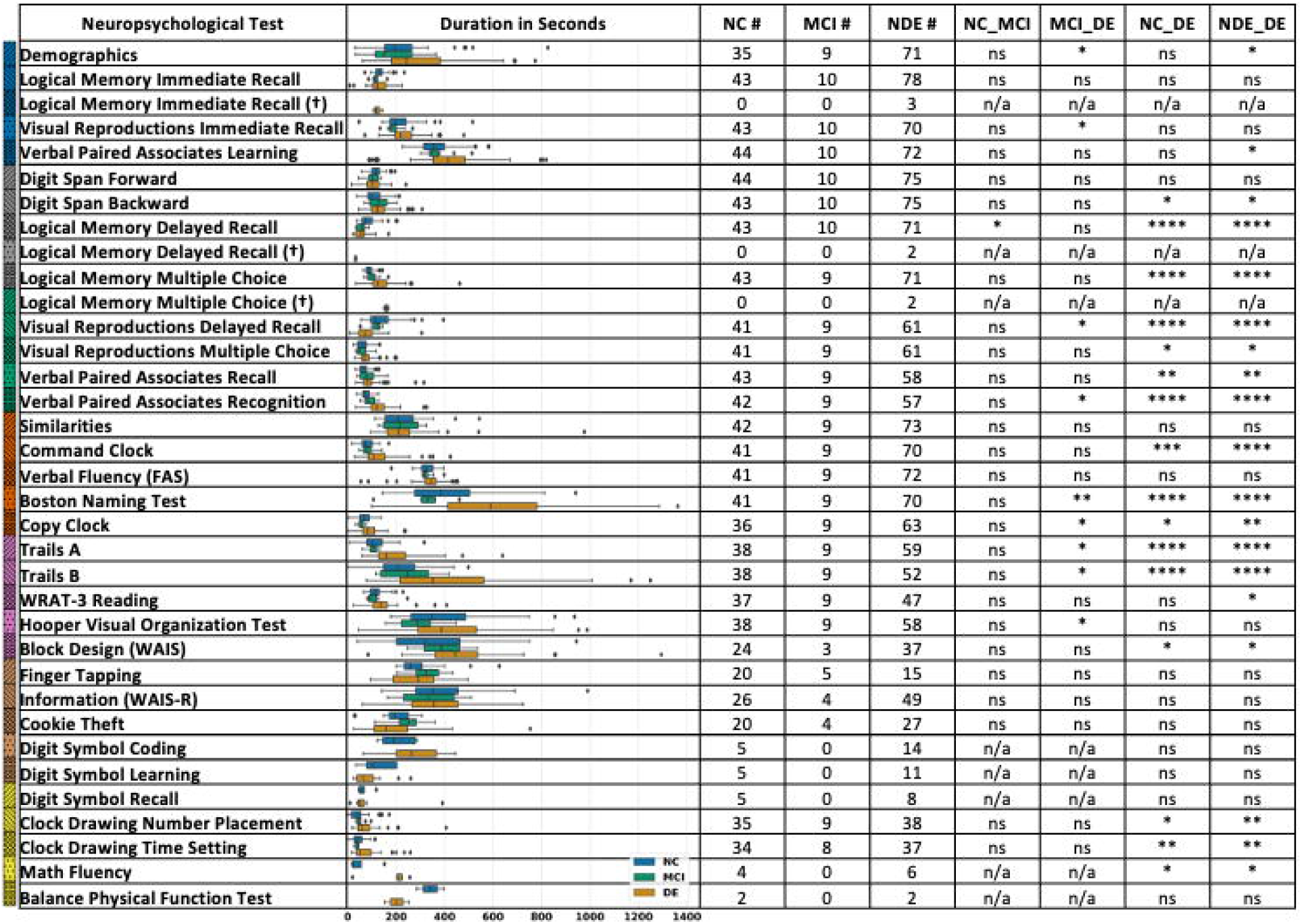
Time spent on the neuropsychological tests. Boxplots showing the time spent by the FHS participants on each neuropsychological test. For each test, the boxplots were generated on participants with normal cognition (NC), those with mild cognitive impairment (MCI), and those who had dementia (DE); those who were non-demented (NDE) combined the NC and MCI individuals. We also indicated the number of recordings that were processed to generate each boxplot. We also computed pairwise statistical significance between two groups (NC vs MCI, MCI vs DE, NC vs DE and DE vs NDE). We evaluated the differences in means of the durations of all three cognitive statuses using a pairwise t-test. The symbol ‘*’ indicates statistical significance at p<0.05, the symbol ‘**’ indicates statistical significance at p<0.01, the symbol ‘***’ indicates statistical significance at p<0.001, and ‘n.s.’ indicates p>0.05. Logical Memory (LM) tests with a (†) symbol denote that an alternative story prompt was administered for the test. It is possible that one participant may receive a prompt under each of the LM recall conditions (one recording). Because many neuropsychological tests were administered on the participants, we chose a representation scheme that combined colors and hatches. The colored hatches were used to represent each individual neuropsychological test and this information was used to aid visualization in subsequent figures.

### Data preprocessing

To preserve as much useful information as possible, the Mel-frequency cepstral coefficients (MFCCs) were extracted during the data preprocessing stage. MFCCs are the coefficients that collectively make up the Mel-frequency cepstrum, which serves as an important acoustic feature in many speech processing applications, particularly in medicine [10-15]. Leveraging the nonlinear Mel scaling makes the response much closer to the human auditory system and therefore renders it an ideal feature for speech-related tasks. Each FHS audio recording was first split into short frames with a window size of 25 ms (i.e., 200 sample points at 8000Hz sampling rate) and a stride length of 10 ms. For each frame, the periodogram estimate of the power spectrum was calculated by 256-point discrete Fourier transformation. Then, a filterbank of 26 triangle filters evenly distributed with respect to the Mel scale was applied to each frame. We then applied a discrete Cosine transformation of the logarithm of all filterbank energies. Note that the 26 filters correspond to 26 coefficients, but in practice, only 2^nd^-13^th^ coefficients are believed to be useful; we loosely followed this convention while replacing the first coefficient with total energy that might contain helpful information on the entire frame.

### Hierarchical long short-term memory network model

Recurrent neural networks (RNN) have long been used for capturing complex patterns in sequential data. Because the durations of the FHS recordings averaged more than one hour and corresponded to hundreds of thousands of MFCCs, a single RNN may not be able to memorize and identify patterns across such long sequences. We therefore developed a two-level hierarchical long short-term memory network (LSTM) model [16], to associate the raw voice recordings with dementia status (**Figure 2A**). Our previous work has shown how the unique architecture of the LSTM model can tackle long sequences of data [17]. Also, as a popular variation in the RNN family, the LSTM model is well suited to overcoming issues such as vanishing gradients and long-term dependencies compared to other RNN frameworks.

**Figure 2:**
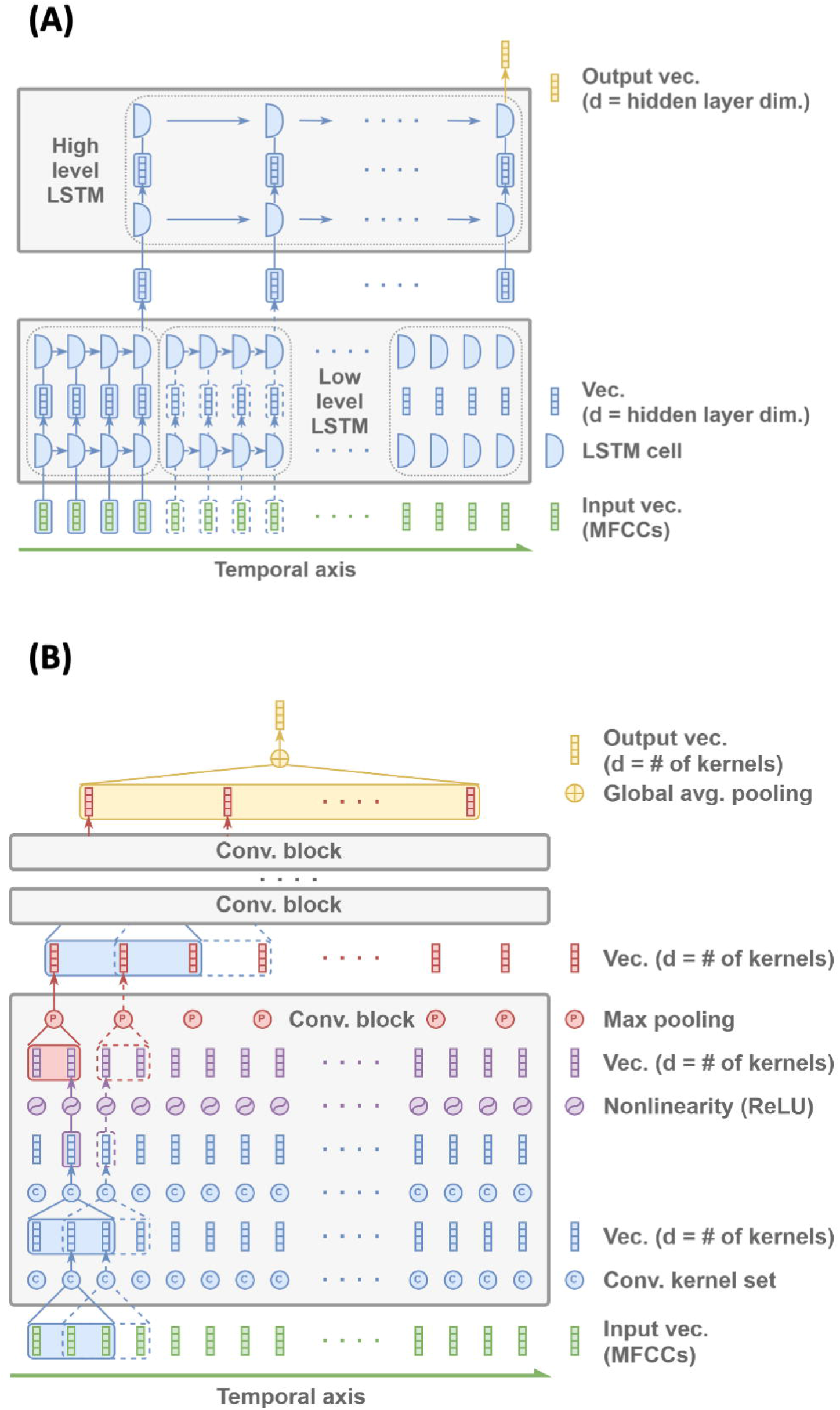
Schematics of the deep learning frameworks. (A) Hierarchical long short-term memory (LSTM) network model that encodes an entire audio file into a single vector to predict dementia status on the individuals. All LSTM cells within the same row share the parameters. Note that the hidden layer dimension is user-defined (e.g., 64 in our approach.) (B) Convolutional neural network that uses the entire audio file as the input to predict the dementia status of the individual. Each convolutional block reduces the input length by a common factor (e.g., 2) while the very top layer aggregates all remaining vectors into one by averaging them.

A one-hour recording may yield hundreds of thousands of temporally ordered MFCCs vectors, while the memory capacity of the LSTM model is empirically limited to only a few thousand vectors. We first grouped every 2000 consecutive MFCC vectors into segments without overlap. For each segment, the low-level LSTM took 10 MFCCs at a time and moved onto the next 10 MFCCs until the end. We then collected the last hidden state as the low-level feature vector for the segment. After processing all those segments one by one, the collection of the low-level feature vectors formed another sequence, which was then fed into the high-level LSTM to generate a high-level feature vector summarizing the entire recording. Note that the hierarchical design ensured that the two-level LSTM architecture was not overwhelmed by longer sequences beyond their practical limitation. For the last step of the LSTM scheme, a multilayer perceptron (MLP) network was used to estimate the probability of dementia based on the summarized vector. Both the low-level and the high-level LSTM shared the same hyperparameters where the hidden state dimension was 64 and the initial states were all set to zeros. The MLP was combined with a 64-dimensional hidden layer and an output layer along with a non-linear activation function. The output layer was then followed by a softmax function to project the results onto the probability space.

### Convolutional neural network model

For comparison with the LSTM model, we designed a one-dimensional convolutional neural network (CNN) model for dementia classification (**Figure 2B**). The stem structure of the CNN model consisted of 7 stacked convolutional blocks. Within each block, there were 2 convolutional layers, 1 max-pooling layer and 1 nonlinearity function. All the convolutional layers had a filter size of 3, stride size of 1 and a padding size of 1. For the pooling layers in the first 6 blocks, we used max pooling with a filter size of 4 and a stride size of 4. The last layer used global average pooling to tackle audio recordings of variable lengths. By applying global average pooling, all the information gets transformed to a fixed-length feature vector, making it straightforward for classification. The CNN stem structure was then followed by a linear classifier which comprised of 1 convolutional layer and 1 softmax layer. Note that the filter size of the convolutional layer was set to be the exact size of the output feature vector.

### Saliency maps

We derived the saliency maps based on the intermediate result right before the global average pooling step of the CNN model. The intermediate result was composed of two vectors which signified DE[+] and DE[-] prediction respectively. For simplicity, we only used the DE[+] vector. Since the recording-level prediction was determined by the average of the saliency map which also preserved temporal structure, it allowed us to observe finer aspects of the prediction by revealing the values assigned to each short period. For our CNN model settings, each value corresponded to roughly two minutes and a half of an original recording. Note that the length of the periods is implied by the CNN model parameters; altering the stride size, the kernel size, the number of stacked CNN blocks etc. may result in different lengths. To align the saliency map with the original recording for further analysis, we extended its size to the total number of seconds via nearest neighbor interpolation.

### Salient administered fractions

The CNN model tasked with classifying between participants with normal cognition and dementia was tested on 81 participants who have transcripts for 123 recordings out of their 183 total recordings. Those 60 recordings without transcripts were excluded from the saliency analysis but were included in the test dataset. Of the recordings with transcripts, there were 44 recordings of participants with normal cognition and 79 recordings of participants with dementia. The recordings that were transcribed were divided into subgroups based on the time spent by the participant on each neuropsychological test. As a result, for each second of a given recording, the DE[+] saliency value and the current neuropsychological test are known. In order to calculate the salient administered fraction (SAF) for a given neuropsychological test, we counted the number of seconds for that neuropsychological test that also had a DE[+] saliency value greater than zero and then divided it by the total number of seconds for that neuropsychological test. For example, if the Boston Naming Test (BNT) has 90 seconds that had DE[+] saliency values greater than zero and BNT was administered for 100 seconds, then the SAF[+] would be 0.90. Similarly, we produced SAF[-] in the same way, except for periods of time where the DE[+] saliency value was less than or equal to zero. The SAF[+] was calculated for every administered neuropsychological test for each recording that had a demented participant and that the model classified the participant as demented (true positive). The SAF[-] was calculated in the same way, except for recordings with participants with normal cognition and the model classified to have normal cognition (true negative).

### Data organization, model performance and statistical analysis

The dataset for this study includes digital voice recordings from September 2005 to March 2020 from the subset of FHS participants who came to dementia review. The models were implemented using PyTorch 1.4 and constructed on a workstation with a GeForce RTX 2080 Ti graphics processing unit. A portion of the participants along with their recordings were kept aside for independent model testing (**Figure S1**). Note that manual transcriptions were available on these cases, which allowed us to perform saliency analysis. Using the remaining data, the models were trained using 5-fold cross-validation. We split the data on the participant level for each fold and then all of a given participant’s recordings were included in each fold. We acknowledge that this does not exactly split the data into even folds because a participant may have a different number of recordings compared to another participant, however each participant generally had a similar distribution of recordings, which mitigated this effect.

We also tested the model performance on audio segments of shorter lengths. From the test data, we randomly extracted 5-minute, 10-minute and 15-minute recordings from the participants and grouped them based on the audio length. Both the LSTM and CNN models trained on the full audio recordings were used to predict on these short audio segments. Note that only one segment (5-min or 10-min or 15-min) was extracted with replacement per recording in one round of testing. This process was repeated five times and results were reported.

We evaluated the differences in means of the durations of all three cognitive statuses using a pairwise t-test, which is available in Python’s SciPy package. Performance of the machine learning models was presented as mean and standard deviation over the model runs. We generated receiver operating characteristic (ROC) and precision-recall (PR) curves based on the cross-validated model predictions. For each ROC and PR curve, we also computed the area under curve (AUC) values. Additionally, we computed model accuracy, balanced accuracy, sensitivity, specificity, F1-score, weighted F1-score and Matthews correlation coefficient on the test data.

## Results

Both the LSTM and the CNN models that were trained and validated on the FHS voice recordings demonstrated consistent performance across the different data splits used for 5-fold cross validation (**Figure 3**). For the classification of demented versus normal participants, the LSTM model took 20 minutes to fully train (8 epochs and batch size of 4) and took 14 seconds to predict on a test case; the CNN model took 106 minutes to fully train (32 epochs and batch size of 4) and took 13 seconds to predict on a test case. For the classification of demented versus non-demented participants, the LSTM model took 187 minutes to fully train (32 epochs and batch size of 4) and took 20 seconds to predict on a test case; the CNN model took 427 minutes to fully train (64 epochs and batch size of 4) and took 20 seconds to predict on a test case. In general, we observed that the CNN model tested on the full recordings performed better on most metrics for the classification problem focused on distinguishing participants with DE from those that have NC (**Tables 3A & 3B, Figure S2**). The only exception was that the LSTM model mean specificity was higher than the mean specificity of the CNN model. For the classification problem focused on distinguishing participants with DE from those who were NDE, both models performed evenly (**Tables 3C & 3D, Figure S3**). The LSTM model’s sensitivity was higher, but the CNN model’s specificity was higher than its counterpart. Interestingly, in some cases, the model performance on the shorter segments was higher than the full audio recordings. For the classification problem focused on distinguishing participants with DE from those that have NC, we found improved LSTM model performance on both 10- and 15-min recordings, based on a few metrics. For the same classification problem, the CNN model’s performance on the full audio recordings mostly the highest, with the 5-min segment’s mean sensitivity being the exception. For the classification problem focused on distinguishing participants with DE from those who were NDE, the LSTM model’s performance on the full recordings was highest based on all the metrics. For the same classification problem, the CNN model’s performance on the full audio recordings mostly the highest, with the 5-min segment’s mean sensitivity and the 10-min mean F1-score being the exceptions.

**Table 3:** Performance of the deep learning models. The long short-term memory (LSTM) network and the convolutional neural network (CNN) models were constructed to classify participants with normal cognition and dementia as well as participants who are non-demented and the ones with dementia, respectively. On each model, a 5-fold cross-validation was performed and the model predictions (mean ± standard deviation) were generated on the test data (see Figure S1). Tables (A) and (B) report the performances of the LSTM and the CNN models for the classification of participants with normal cognition versus those with dementia. Tables (C) and (D) report the performances of the LSTM and the CNN models for the classification of participants who are non-demented versus those who have dementia.

| **Normal vs. Demented Classification (LSTM model)** |  |  |  |  |
| --- | --- | --- | --- | --- |
| **Model** | **LSTM-5 minutes** | **LSTM-10 minutes** | **LSTM-15 minutes** | **LSTM-Full audio** |
| **Accuracy** | 0.581±0.039 | 0.578±0.037 | 0.593±0.051 | **0.598±0.035** |
| **Balanced Accuracy** | 0.642±0.029 | 0.641±0.027 | **0.650±0.035** | 0.647±0.027 |
| **Sensitivity** | 0.420±0.065 | 0.412±0.067 | 0.442±0.093 | **0.470±0.077** |
| **Specificity** | 0.865±0.022 | **0.871±0.031** | 0.859±0.034 | 0.824±0.025 |
| **Precision** | 0.844±0.019 | **0.849±0.029** | 0.846±0.025 | 0.824±0.010 |
| **F1 Score** | 0.558±0.061 | 0.551±0.062 | 0.575±0.083 | **0.595±0.063** |
| **Weighted F1 Score** | 0.573±0.046 | 0.569±0.046 | 0.586±0.061 | **0.596±0.047** |
| **MCC** | 0.294±0.050 | 0.294±0.049 | **0.307±0.060** | 0.294±0.046 |
| **Precision-Recall AUC** | 0.814±0.016 | **0.819±0.020** | 0.803±0.029 | 0.805±0.022 |
| **ROC AUC** | 0.742±0.017 | **0.745±0.011** | 0.737±0.020 | 0.740±0.017 |

**(B)**
| **Normal vs. Demented Classification (CNN model)** |  |  |  |  |
| --- | --- | --- | --- | --- |
| **Model** | **CNN-5 minutes** | **CNN-10 minutes** | **CNN-15 minutes** | **CNN-Full audio** |
| **Accuracy** | 0.666±0.035 | 0.674±0.052 | 0.710±0.021 | **0.740±0.033** |
| **Balanced Accuracy** | 0.587±0.054 | 0.650±0.035 | 0.698±0.015 | **0.743±0.015** |
| **Sensitivity** | **0.873±0.079** | 0.738±0.118 | 0.740±0.045 | 0.735±0.094 |
| **Specificity** | 0.300±0.160 | 0.562±0.095 | 0.656±0.038 | **0.750±0.083** |
| **Precision** | 0.691±0.036 | 0.750±0.025 | 0.792±0.013 | **0.844±0.034** |
| **F1 Score** | 0.769±0.028 | 0.738±0.064 | 0.765±0.023 | **0.780±0.048** |
| **Weighted F1 Score** | 0.623±0.061 | 0.672±0.047 | 0.712±0.019 | **0.742±0.033** |
| **MCC** | 0.207±0.106 | 0.308±0.077 | 0.389±0.034 | **0.477±0.026** |
| **Precision-Recall AUC** | 0.743±0.038 | 0.801±0.024 | 0.837±0.012 | **0.876±0.028** |
| **ROC AUC** | 0.640±0.054 | 0.716±0.038 | 0.759±0.019 | **0.805±0.027** |

(C)
| **Nondemented vs. Demented Classification (LSTM model)** |  |  |  |  |
| --- | --- | --- | --- | --- |
| **Model** | **LSTM-5 minutes** | **LSTM-10 minutes** | **LSTM-15 minutes** | **LSTM-Full audio** |
| **Accuracy** | 0.651±0.016 | 0.659±0.022 | 0.648±0.023 | **0.675±0.013** |
| **Balanced Accuracy** | 0.651±0.016 | 0.659±0.022 | 0.648±0.023 | **0.675±0.013** |
| **Sensitivity** | 0.576±0.048 | 0.565±0.062 | 0.556±0.059 | **0.578±0.049** |
| **Specificity** | 0.726±0.031 | 0.753±0.024 | 0.740±0.035 | **0.772±0.027** |
| **Precision** | 0.677±0.016 | 0.694±0.012 | 0.680±0.025 | **0.716±0.011** |
| **F1 Score** | 0.621±0.027 | 0.621±0.040 | 0.610±0.038 | **0.638±0.028** |
| **Weighted F1 Score** | 0.649±0.016 | 0.655±0.024 | 0.644±0.025 | **0.671±0.015** |
| **MCC** | 0.306±0.031 | 0.324±0.040 | 0.302±0.046 | **0.357±0.022** |
| **Precision-Recall AUC** | 0.685±0.012 | 0.682±0.019 | 0.670±0.025 | **0.701±0.016** |
| **ROC AUC** | 0.720±0.013 | 0.726±0.009 | 0.711±0.019 | **0.734±0.014** |

(D)
| **Nondemented vs. Demented Classification (CNN model)** |  |  |  |  |
| --- | --- | --- | --- | --- |
| **Model** | **CNN-5 minutes** | **CNN-10 minutes** | **CNN-15 minutes** | **CNN-Full audio** |
| **Accuracy** | 0.555±0.022 | 0.624±0.030 | 0.628±0.042 | **0.653±0.020** |
| **Balanced Accuracy** | 0.555±0.023 | 0.623±0.030 | 0.627±0.042 | **0.652±0.020** |
| **Sensitivity** | **0.663±0.224** | 0.546±0.101 | 0.486±0.076 | 0.457±0.106 |
| **Specificity** | 0.447±0.188 | 0.701±0.065 | 0.769±0.038 | **0.847±0.068** |
| **Precision** | 0.543±0.011 | 0.646±0.034 | 0.674±0.053 | **0.760±0.049** |
| **F1 Score** | 0.576±0.120 | **0.587±0.055** | 0.563±0.063 | 0.560±0.068 |
| **Weighted F1 Score** | 0.528±0.035 | 0.619±0.030 | 0.619±0.045 | **0.635±0.031** |
| **MCC** | 0.128±0.055 | 0.253±0.062 | 0.265±0.085 | **0.337±0.024** |
| **Precision-Recall AUC** | 0.597±0.041 | 0.643±0.033 | 0.655±0.044 | **0.732±0.015** |
| **ROC AUC** | 0.595±0.043 | 0.663±0.033 | 0.683±0.037 | **0.746±0.021** |

**Figure 3:**
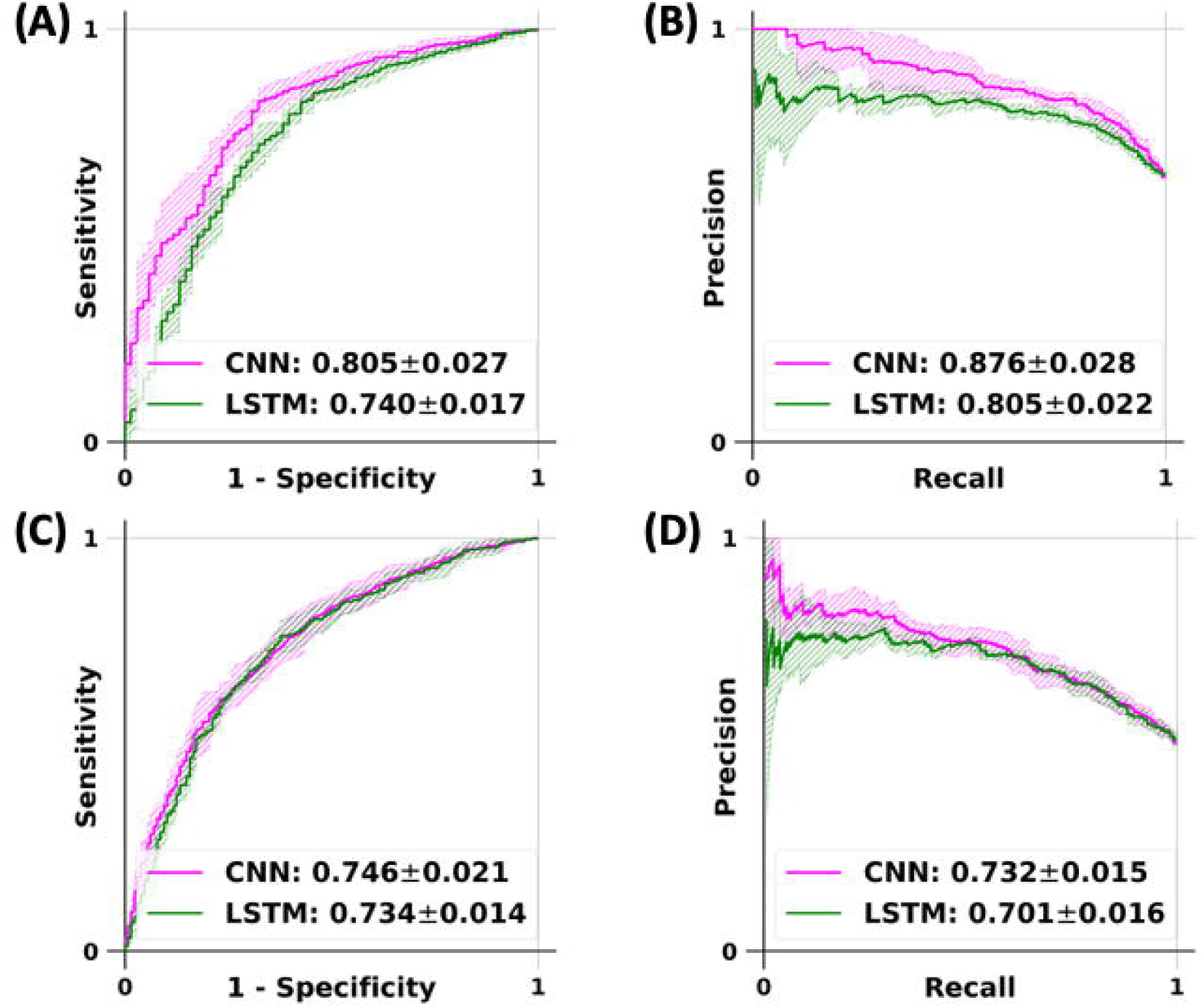
Receiver operating characteristic (ROC) and precision-recall (PR) curves of the deep learning models. The long short-term memory (LSTM) network and the convolutional neural network (CNN) models were constructed to classify participants with normal cognition and dementia as well as participants who are non-demented and the ones with dementia, respectively. On each model, a 5-fold cross-validation was performed and the model predictions (mean ± standard deviation) were generated on the test data (see Figure S1), followed by creation of the ROC and PR curves. Plots (A) and (B) denote the ROC and PR curves for the LSTM and the CNN models for the classification of normal versus demented cases. Plots (C) and (D) denote the ROC and PR curves for the LSTM and CNN models for the classification of non-demented versus demented cases.

We computed the average (± standard deviation) of SAF[+] and SAF[-] derived from the CNN model (**Table 4**). The positive SAFs were calculated for all true positive recordings and the negative SAFs were calculated for all true negative recordings. For example, the SAF[+] for the ‘Verbal paired associates recognition’ test was 0.88±0.26. This indicates that on average 88% of the duration of the ‘Verbal paired associates recognition’ tests administered in true positive recordings also intersected with segments of time that the model found to be DE[+] salient. Also, the SAF[-] for the ‘Verbal paired associates recognition’ test was 0.32±0.42. This indicates that on average 32% of the duration of the ‘Verbal paired associates recognition’ tests administered in true negative recordings also intersected with segments of time that the model found to not be DE[+] salient. On the other hand, the SAF[+] for the ‘Command clock’ test was 0.39±0.45, indicating that only about 39% of the duration of the “Command clock’ tests administered in true positive recordings also intersected with segments of time that the model found to be DE[+] salient. Also, the SAF[-] for the ‘Command clock’ test was 0.76±0.37. The rest of the average positive SAFs and average negative SAFs were also reported for the remaining neuropsychological tests as well as the number of true positive or true negative recordings that contained an administration of the given test (**Table 4**). We also computed average SAF[+] and SAF[-] for additional neuropsychological tests (**Table S2)**, which were set aside due to the low number of samples. A schematic of the DE[+] saliency vectors used to generate SAF[+] and SAF[-] can be seen in **Figure 4**. Each value in the saliency vector represented approximately 2 minutes and 30 seconds of a recording. Since the saliency vector covers the entire recording, each second of every neuropsychological test within a recording can be assigned a saliency vector value, which was then used to calculate SAF[+] and SAF[-].

**Table 4:** Salient administered fractions derived from the CNN model. The average salient administered fraction (SAF) and standard deviation for true positive (SAF[+]) and true negative (SAF[-]) cases are listed in descending order based on the SAF[+] value. SAF[+] is calculated by summing up the time spent in a given neuropsychological test that intersects with a segment of time that is DE[+] salient and dividing by the total time spent in a given neuropsychological test. SAF[-] is calculated by summing up the time spent in a given neuropsychological test that intersects with a segment of time that is not DE[+] salient and dividing by the total time spent in a given neuropsychological test. The number of samples for SAF[+] and SAF[-] indicate the number of true positive and true negative recordings that contain each neuropsychological test.

| Test | SAF[+] | SAF[-] | SAF[+] samples | SAF[-] samples |
| --- | --- | --- | --- | --- |
| **Verbal paired associates recognition** | 0.88±0.26 | 0.32±0.42 | 37 | 32 |
| **Verbal paired associates learning** | 0.83±0.22 | 0.23±0.22 | 51 | 33 |
| **Boston naming test** | 0.82±0.18 | 0.30±0.29 | 47 | 31 |
| **Digit span forward** | 0.78±0.37 | 0.16±0.33 | 53 | 33 |
| **Wrat-3 reading** | 0.76±0.36 | 0.57±0.43 | 30 | 27 |
| **Information (WAIS-R)** | 0.73±0.28 | 0.36±0.30 | 36 | 16 |
| **Verbal paired associates recall** | 0.72±0.41 | 0.33±0.41 | 38 | 32 |
| **Finger tapping** | 0.71±0.33 | 0.47±0.33 | 11 | 15 |
| **Digit span backward** | 0.70±0.42 | 0.36±0.43 | 53 | 32 |
| **Demographics** | 0.69±0.33 | 0.22±0.35 | 55 | 25 |
| **Trails B** | 0.68±0.30 | 0.65±0.35 | 34 | 28 |
| **Logical memory immediate recall** | 0.66±0.42 | 0.72±0.40 | 56 | 32 |
| **Verbal fluency (FAS)** | 0.66±0.34 | 0.39±0.27 | 49 | 31 |
| **Similarities** | 0.66±0.38 | 0.56±0.34 | 50 | 32 |
| **Clock drawing time setting** | 0.66±0.48 | 0.48±0.51 | 24 | 25 |
| **Logical memory delayed recall** | 0.65±0.44 | 0.74±0.36 | 50 | 32 |
| **Cookie theft** | 0.64±0.41 | 0.65±0.39 | 19 | 14 |
| **Block design (WAIS)** | 0.64±0.27 | 0.70±0.33 | 26 | 14 |
| **Clock drawing number placement** | 0.63±0.45 | 0.49±0.49 | 26 | 26 |
| **Visual reproductions multiple choice** | 0.60±0.47 | 0.44±0.48 | 41 | 30 |
| **Hooper visual organization test** | 0.58±0.29 | 0.48±0.26 | 39 | 28 |
| **Logical memory multiple choice** | 0.57±0.42 | 0.87±0.28 | 50 | 32 |
| **Visual reproductions immediate recall** | 0.55±0.35 | 0.85±0.21 | 48 | 32 |
| **Trails A** | 0.52±0.41 | 0.51±0.44 | 40 | 28 |
| **Visual reproductions delayed recall** | 0.52±0.47 | 0.77±0.33 | 40 | 30 |
| **Copy clock** | 0.44±0.45 | 0.36±0.47 | 41 | 26 |
| **Command clock** | 0.39±0.45 | 0.76±0.37 | 47 | 31 |

**Figure 4:**
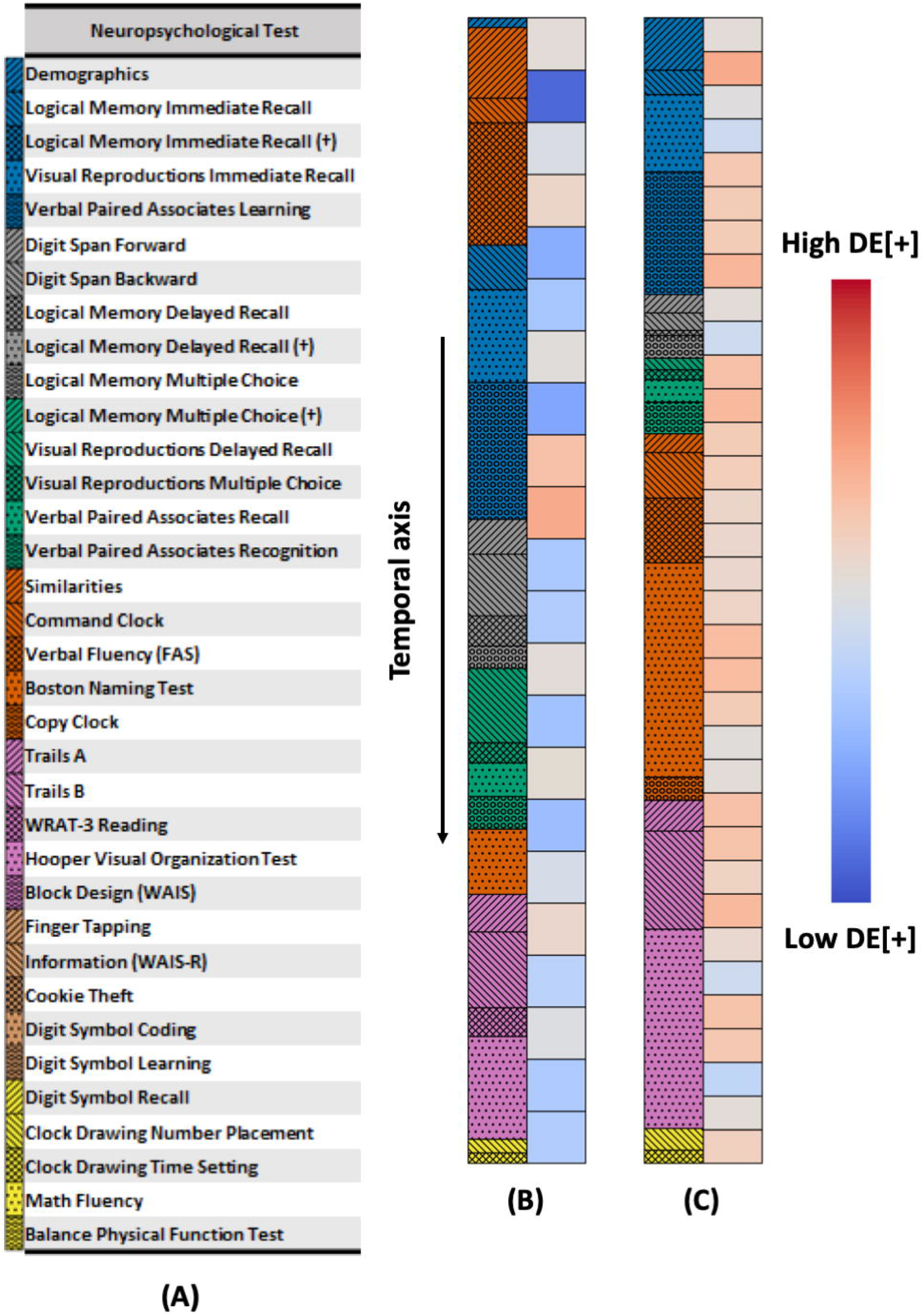
Saliency maps highlighted by the CNN model. (A) This key is a representation that maps the colored hatches to the neuropsychological tests. (B) Saliency map representing a recording (62 minutes in duration) of a participant with normal cognition (NC) that was classified as NC by the convolutional neural network (CNN) model. (C) Saliency map representing a recording (94 minutes in duration) of a participant with dementia (DE) who was classified with dementia by the CNN model. For both (B) & (C), the colormap on the left half corresponds to a neuropsychological test. The color on the right half represents the DE[+] value, ranging from dark blue (low DE[+]) to dark red (high DE[+]). Each DE[+] rectangle represents roughly 2 minutes and 30 seconds.

## Discussion

Cognitive performance is affected by numerous factors that are independent of underlying actual cognitive status. Physical impairments (vision, hearing), mood (depression, anxiety), low education, cultural bias, as well as test-specific characteristics are just a few of many variables that can lead to variable performance. Further, neuropsychological tests that claim to test the same specific cognitive domains (e.g., memory, executive function, language, etc.) do not do so in a uniform manner. For example, when testing verbal memory, a paragraph recall test taps into a different set of underlying cognitive processes compared to that of a list learning task. Also, the presumption that a test measures only a single cognitive domain is naively simplistic since every neuropsychological test involves multiple cognitive domains to complete. Thus, across many factors, there is significant heterogeneity of cognitive performance that make it difficult to differentiate between those who will become demented versus those who will not, especially at the preclinical stage. Particularly, traditional paper and pencil neuropsychological tests may not be sufficiently sensitive to pick up when subtle change begins. While cognitive complaints serve as a surrogate preclinical measure of decline, there is the inherent bias of self-report. Given this complex landscape, digital voice recordings of neuropsychological tests provide a data source of relative independence from the limitations. To our knowledge, our study is the first to demonstrate that a continuous stream of data is also amenable for automated analysis for evaluation of cognitive status.

Digital health technologies in general and voice in particular, are increasingly being evaluated as potential screening tools for depression [18-21], and various neurodegenerative diseases such as Parkinson’s disease [22-25]. Recently, potential opportunities for developing digital biomarkers based on mobile/wearables for AD were outlined [26, 27]. Our study is unique in focusing on two deep learning methods that relies on a completely hands-free approach for processing raw voice recordings to predict dementia status. The advantage of our approach is three-fold. The first is the limited need to extensively process any of the voice recordings before sending them as inputs to the neural networks. This is a major advantage because it minimizes the burden of generating accurate transcriptions and/or handcrafted features that generally take time to develop and relying on the availability of experts who are not necessarily readily available. This aspect places us in a unique position compared to previously published work that depended on derived measures [28, 29]. Second, our approach can process audio recordings of variable lengths, which means that one does not have to format or select a specific window of the audio recording for analysis. This important strength underscores the generalizability of our work because one can process voice recordings containing various combinations of neuropsychological tests that are not bounded within a time frame. Finally, our approach allows for identification of audio segments that are highly correlated with the outcome of interest. The advantage of doing this is that it provides a “window” into the machine learning black box; we can go back to the recordings and identify the various speech patterns or segments of the neuropsychological tests, which point to high probability of disease risk and understand their contextual significance.

The CNN architecture allowed us to generate the saliency vectors by utilizing the parameters of the final classification layer. Simply put, a temporal saliency vector for each specific case could be obtained by calculating the weighted sum of the output feature vectors from the last convolutional block in the CNN, indicating how each portion of the recording contributed to either positive or negative prediction. We then aligned the saliency vectors with the recording timeline to further analyze the speech signatures to understand if there were any snippets of the neuropsychological testing that often were correlated with the output class label. From examining the transcriptions that exist for a portion of the dataset, we were able to identify which neuropsychological tests were occurring during any given time in a recording, and then calculated the positive SAFs. This implies that the neuropsychological tests found in these segments may be presenting a test in which the participant’s voice in their response has a signal related to their cognition. For example, the SAF[+] is high for the “Verbal paired associates recognition” test, which could mean that the participants’ audio signals during this test highly influenced the model performance. This result could also imply that most participants diagnosed with dementia may have had explicit episodic memory deficits. The exact connections between the voice signals in the segments identified by the saliency maps and their clinical relevance is worth exploring in the future.

All the FHS voice recordings contained audio content from two distinct speakers of which one is the interviewee (participant), and the other is the interviewer (clinician). We did not attempt to discern speaker-specific audio content as our models processed the entire raw audio recording at once. This choice was intentional because we wanted to first evaluate if deep learning can predict dementia status of the participant without having to perform detailed feature engineering on the audio recordings. Future work could focus on processing these signals and recognizing speaker-related differences and interactions, periods of silence and other nuances to make the audio recordings more amenable for deep learning. Also, additional studies can be performed to integrate the audio data with other routinely collected information that requires no additional processing (i.e., demographics) to augment model performance. An important point to note is that studies as proposed above need to be conducted with the goal of creating scalable solutions across the globe, particularly to those regions where technical or advanced clinical expertise is not readily available. This means that users at the point-of-care may not be in the best position to manually process raw voice recordings or any other data to derive needed features that can be fed into the computer models. Since our deep learning models do not require data pre-processing or handcrafted features, our approach can serve as a potential screening tool without having the need of an expert-level input. Our current findings serve as a first step towards achieving such a solution that can have a broader outreach.

### Study limitations

The models were developed using the FHS voice recordings, which is a single population cohort from the New England area in the United States. Despite demonstrating consistent model performance using rigorous cross-validation (5-fold) approaches, our models still need to be validated using data from external cohorts to confirm their generalizability. Currently, we do not have access to any other cohort that has voice recordings of neuropsychological exams. Therefore, our study findings need to be interpreted considering this limitation and with the hope of evaluating them further in the future. Due to lack of available data in some cases and because not all participants took all the types of neuropsychological tests, we were able to generate the distribution of times spent for a portion of the tests. It must be also noted that the number of NC, MCI, DE and NDE participants varied for each neuropsychological test. Additionally, there may be outside factors affecting the amount of time it took a participant to complete a neuropsychological test that are not represented in the boxplots. For example, a normal participant could finish the BNT quickly and perform well, whereas administration of the BNT to a demented participant could be abruptly stopped due to an inability to complete the test or for other reasons. Therefore, the amount of time spent administering the BNT in the recordings in those two cases could be similar, but for different reasons. Nevertheless, statistical tests were performed to quantify the pairwise differences on all the available neuropsychological exams, which gave us the flexibility to report the differences that were statistically different and those that were similar. Also, while it is possible that the interviewer’s behavior can influence the interviewee’s response, we must acknowledge that all the interviewers are professionally trained to uniformly administer the neuropsychological tests.

## Conclusions

Our proposed deep learning approaches (LSTM & CNN) to processing raw voice recordings in an automated fashion allowed us to classify dementia status. Such approaches that rely minimally on neuropsychological expertise, audio transcription or manual feature engineering can pave the way towards the development of real-time screening tools in dementia care, especially in resource-limited settings.

## Data Availability

All results, code and documentation will be made available on GitHub (https://github.com/vkola-lab/azrt2021) upon manuscript publication. Data in this study cannot be shared publicly due to regulations of local ethical committees. Data might be made available to researchers who meet the criteria (to be provided once all data are available) for access to confidential data and upon IRB approval.

## Declarations

### Ethics approval and consent to participate

No ethics approval or participant consent was obtained because this study was based on retrospective data.

### Consent for publication

All authors have approved the manuscript for publication

### Availability of data and material

Python scripts and sample data are made available on GitHub (https://github.com/vkola-lab/azrt2021). Data in this study cannot be shared publicly due to regulations of local ethical committees. Data might be made available to researchers upon request. All requests will be evaluated based on institutional and departmental policies.

## Acknowledgments

None

## Author contributions

CX contributed to conceptualization, formal analysis, investigation, methods, validation, and visualization. CK contributed to data curation and verification, formal analysis, investigation, methods, validation, and visualization. IP contributed to conceptualization and validation. RA contributed to the conceptualization, data curation and verification, validation, and supervision. VBK contributed to overall supervision, conceptualization, investigation, validation, visualization, and writing the original and the revised draft. All authors contributed to writing the article and editing and have approved the final manuscript. All authors had full access to all the data in the study and had final responsibility for the decision to submit for publication.

## Supplementary figure and table captions

**Table S1:** Demographics of the participants who were non-demented (NDE; i.e., individuals with normal cognition (NC) and mild cognitive impairment (MCI)) at the time of the voice recordings. ApoE data was unavailable for one Gen 1 participant, thirteen Gen 2 participants, and one New Offspring Cohort (NOS) participant; MMSE data was not collected for all Gen 3, OmniGen 2, and NOS participants.

| Non-demented cases |  |  |  |  |  |  |
| --- | --- | --- | --- | --- | --- | --- |
| Cohort | N | Female | ApoE4+ | Recordings | Age (years) | Mean MMSE |
| Gen 1 | 89 | 60 | 15 | 160 | 91.2±3.1 | 26.7±2.3 |
| Gen 2 | 398 | 201 | 89 | 745 | 76.5±7.8 | 27.6±2.1 |
| Gen 3 | 5 | 1 | 0 | 8 | 60.4±10.6 | NA |
| OmniGen 1 | 12 | 3 | 3 | 16 | 72.2±8.1 | 26.0±2.5 |
| OmniGen 2 | 1 | 1 | 1 | 1 | 74.0±0.0 | NA |
| NOS | 2 | 1 | 0 | 4 | 86.5±5.3 | NA |
| Total | 507 | 267 | 108 | 934 | 78.8±9.3 | 27.4±2.2 |

**Figure S1:**
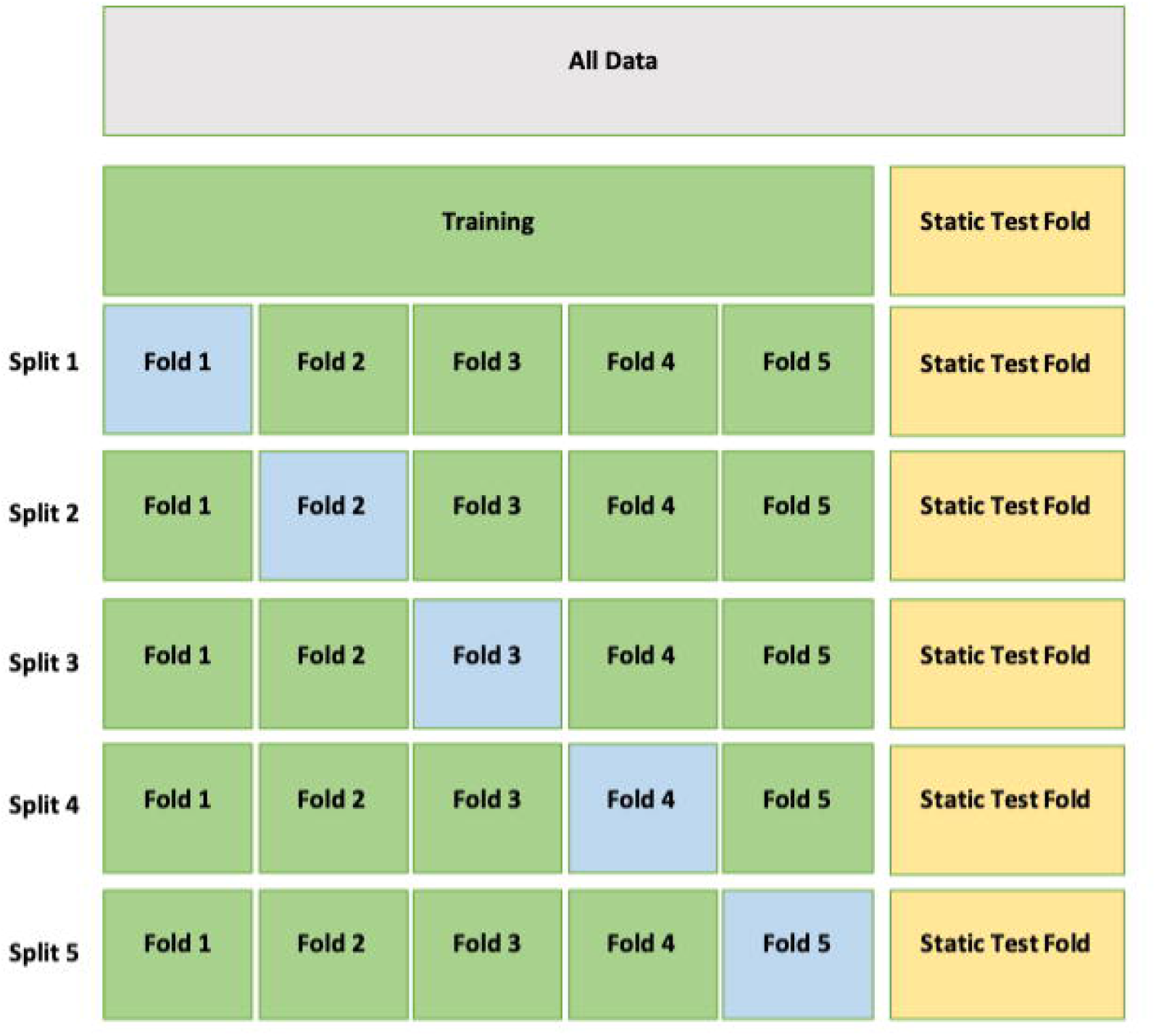
The dataset was first split into two parts such that a portion of the participants along with their recordings were kept aside for independent model testing. The models were trained on the remaining data using 5-fold cross-validation. We split the data on the participant level for each fold and then all of a given participant’s recordings were included in each fold.

**Figure S2:**
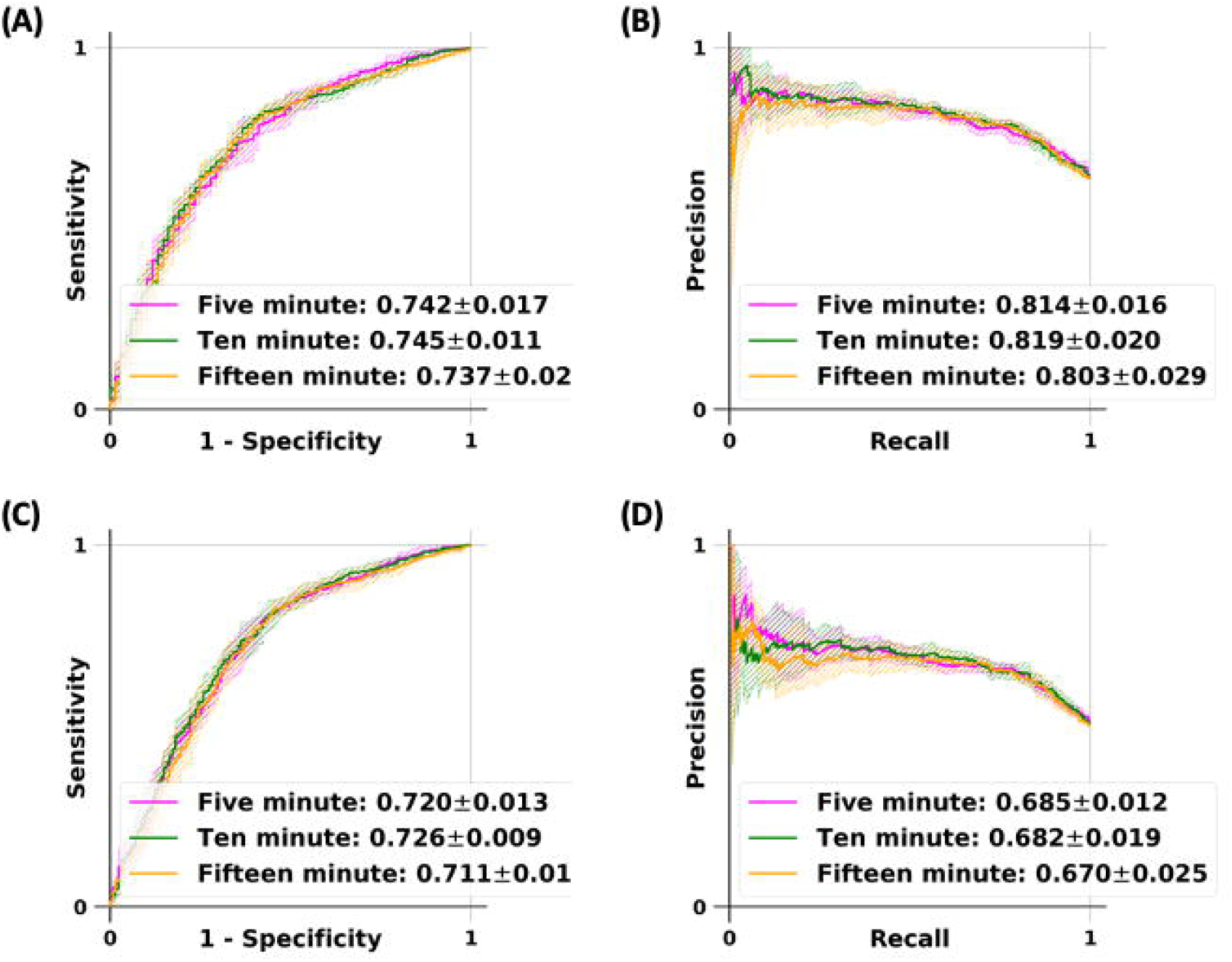
The long short-term memory (LSTM) network and the convolutional neural network (CNN) models were used to classify participants who have normal cognition from those with dementia. Models were trained on the full dataset and the performance was reported on audio samples of variable lengths extracted from the test data (see Figure S1). Plots (A) and (B) denote the ROC and PR curves for the LSTM model and plots (C) and (D) denote the ROC and PR curves for the CNN model.

**Figure S3:**
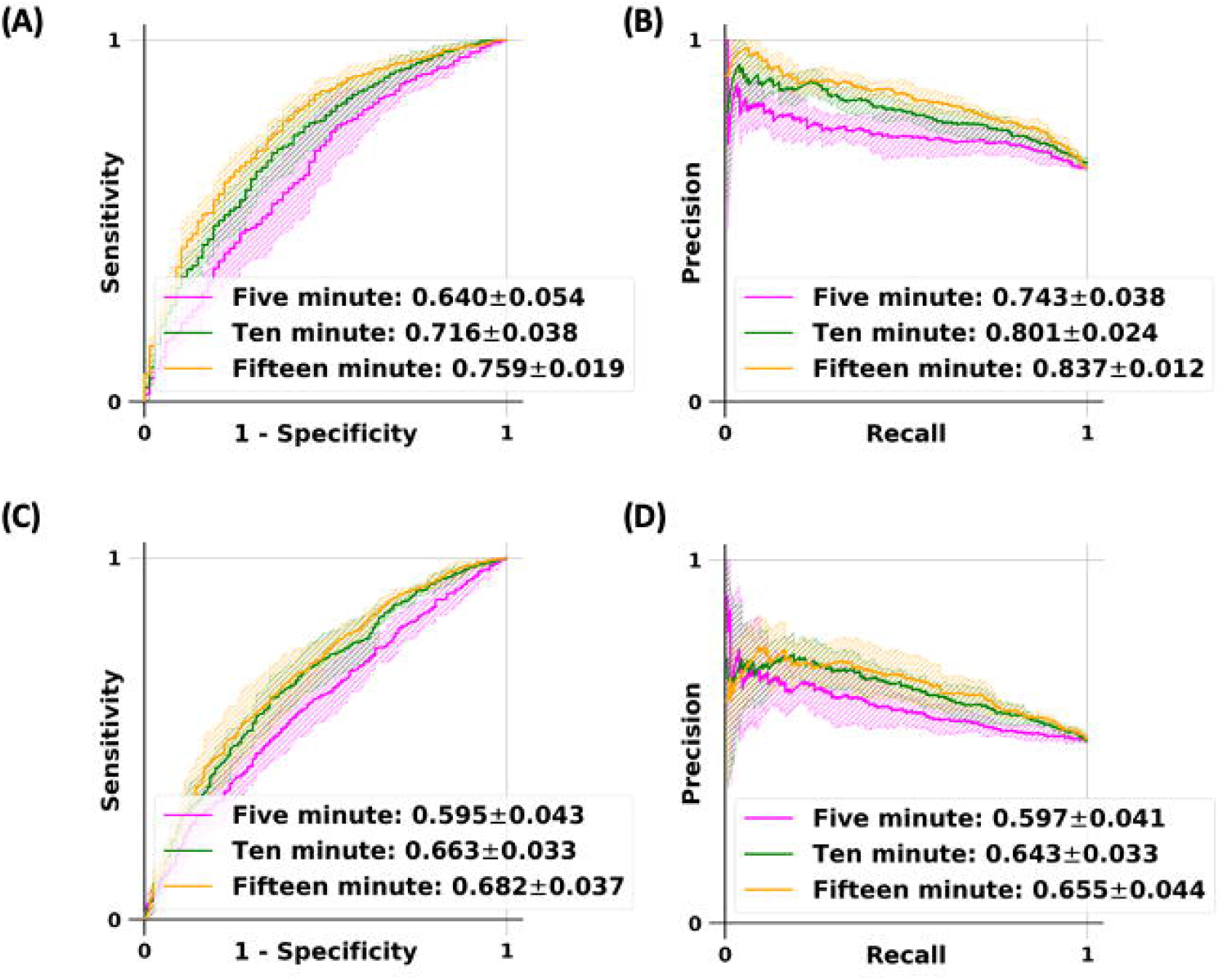
The long short-term memory (LSTM) network and the convolutional neural network (CNN) models were used to classify participants who were not demented from those with dementia. Models were trained on the full dataset and the performance was reported on audio samples of variable lengths extracted from the test data (see Figure S1). Plots (A) and (B) denote the ROC and PR curves for the LSTM model and plots (C) and (D) denote the ROC and PR curves for the CNN model.

**Table S2:** For the neuropsychological tests that have too few samples and for the labeled segments that were unrelated to neuropsychological testing (Miscellaneous and Unlabeled), the average SAF (salient administered fraction) and standard deviation for true positive (SAF[+]) and true negative (SAF[-]) cases are listed in descending order based on the SAF[+] value. SAF[+] is calculated by summing up the time spent in a given neuropsychological test that intersects with a segment of time that is DE[+] salient and dividing by the total time spent in a given neuropsychological test. SAF[-] is calculated by summing up the time spent in a given neuropsychological test that intersects with a segment of time that is not DE[+] salient and dividing by the total time spent in a given neuropsychological test. The number of samples for SAF[+] and SAF[-] indicate the number of true positive and true negative recordings that contain each neuropsychological test. Segments of time were marked as “Miscellaneous” when there was speech in between neuropsychological tests that was unrelated to testing. Segments of time at the start of recordings up until speech occurred were considered “Unlabeled”.

| **Test** | **SAF[+]** | **SAF[-]** | **SAF[+] samples** | **SAF[-] samples** |
| --- | --- | --- | --- | --- |
| **Balance Physical Function Test** | 1.00±0.00 | 0.63±0.30 | 2 | 2 |
| **Logical Memory Delayed Recall (alternate prompt)** | 1.00±0.00 |  | 1 | 0 |
| **Digit Symbol Learning** | 0.77±0.41 | 0.81±0.33 | 6 | 3 |
| **Math Fluency** | 0.74±0.34 |  | 4 | 0 |
| **Digit Symbol Recall** | 0.74±0.46 | 0.40±0.53 | 3 | 3 |
| **Logical Memory Multiple Choice (alternate prompt)** | 0.64±0.00 |  | 1 | 0 |
| **Unlabeled** | 0.61±0.50 | 0.25±0.44 | 31 | 20 |
| **Miscellaneous** | 0.57±0.39 | 0.47±0.45 | 36 | 15 |
| **Logical Memory Immediate Recall (alternate prompt)** | 0.46±0.18 |  | 2 | 0 |
| **Digit Symbol Coding** | 0.43±0.43 | 1.00±0.00 | 9 | 3 |

